# Understanding community knowledge, attitudes and practices related to participation in household transmission investigations during infectious disease outbreaks

**DOI:** 10.64898/2026.04.08.26350464

**Authors:** Niamh Meagher, Dilanka Hettiarachchi, Megan R. Hawkins, Sarah Tavlian, Violeta Spirkoska, Jodie McVernon, Kylie S. Carville, David J. Price, Juan Pablo Villanueva Cabezas, Adrian J. Marcato

## Abstract

**Background:** The World Health Organization has developed several global template protocols for epidemiological investigations, including for household transmission investigations (HHTIs). These investigations facilitate rapid characterisation of novel or re-emerging respiratory pathogens and support evidence-based public health actions. Beyond technical readiness, community buy-in is central to the feasibility and acceptability of HHTIs. Research is needed to determine the perceived legitimacy among the community to inform local protocol adaptation and development of implementation plans that consider community attitudes and needs.

**Methods:** In 2025, we conducted a convenience survey of community members living in Victoria, Australia to explore: their understanding of emerging respiratory diseases; their willingness to take part in public health surveillance activities such as HHTIs; the acceptability of clinical and epidemiological data collection and respiratory/blood sample collection as main components of HHTIs, and; participant comfort towards including their companion animals in HHTIs.

**Results:** We received 282 survey responses, of which 235 were included in the analysis dataset. Compared to the general Victorian population, our participants included a higher proportion of participants who reported being female, tertiary-educated, of Aboriginal and/or Torres Strait Islander heritage, born in Australia and speaking only English at home. Participants indicated overall high levels of comfort and acceptability towards participation in HHTIs, particularly in relation to clinical and epidemiological data collection, with lesser but still high levels of comfort with providing multiple respiratory specimens in a 14-day period. Participants were least comfortable with other specimens such as urine and blood. Involving companion animals in HHTIs was similarly acceptable as human-focused components.

**Conclusions:** Despite our survey population being non-representative of the general Victorian population, our findings provide valuable descriptive insights into the acceptability of HHTIs in Victoria, Australia from which to benchmark future local and international surveys and community engagement activities.

## Background

Enhanced surveillance approaches are essential in outbreaks of novel or re-emerging pathogens to comprehensively conduct risk assessment and inform public health policy decisions.(1) Pathogen transmissibility and severity directly inform response options and the necessary scale of intervention required, including active case follow-up, contact tracing, and potentially disruptive population-wide measures such as social distancing, mobility restrictions and vaccination mandates.

The World Health Organization’s (WHO) *Unity Studies* are global protocol templates for epidemiological investigations that were developed to facilitate rapid characterisation of novel or re-emerging respiratory pathogens to support evidence-based public health action.(2) As part of the *Unity Studies* suite of protocols, the Household Transmission Investigations (HHTIs) protocol is designed to investigate pathogen spread within households, (3) given they are typically closed, well-defined settings where substantial interaction between members leads to high risk of exposure and secondary infection.(4, 5) HHTIs with repeated and regular data and specimen collection can identify infections in household contacts irrespective of symptoms, enabling accurate assessment of pathogens, including transmission dynamics (e.g., secondary attack rate, serial intervals, viral shedding) and infection-severity (e.g., asymptomatic proportion and infection-hospitalisation rate) within the household setting. These characteristics are critical when designing public health policy to limit pathogen spread and mitigate health impacts. Recent outbreaks of mpox, avian influenza and COVID-19 have highlighted the potential for zoonotic and reverse-zoonotic transmission between humans and animals in households and other closed settings,(6-8) leading to calls for expansion of traditional HHTIs to include companion animals as an integrated One Health disease surveillance platform.(9)

Several international COVID-19 response reviews have highlighted the need for improved preparedness for future infectious disease emergencies.(5, 10, 11) While the public availability of *Unity Studies* global template protocols supports readiness and implementation of HHTIs during an outbreak, they are insufficient on their own without detailed local implementation plans.(11) This was evident in Australia’s HHTI for COVID-19 in early-2020. As this was the first study of its kind, the lack of pre-determined and pre-tested implementation plans including considerations for ethics, governance and logistics led to significant delays in study conduct despite buy-in from national committees and health departments. As a result, the study could not generate timely information to support national decision making.(12) A formal study evaluation determined that future local preparedness planning should better integrate HHTIs into core public health surveillance activities and establish functional and adaptable protocols to support rapid activation in outbreaks.(13)

Beyond technical readiness (i.e., availability of protocols, detailed implementation plans, and workforce capability), community involvement is fundamental for pandemic preparedness and response planning,(14) including in the design of enhanced epidemiological investigations such as HHTIs. In the absence of robust literature and evidence on acceptability, positive public attitudes should not be assumed. Research is needed to explore acceptability and perceived legitimacy among the community. Not only will these insights inform feasibility of HHTIs during disease outbreaks, they will also help to shape related preparedness activities that appropriately consider community priorities, preferences and needs. This is particularly important in the post-COVID-19 context, where misinformation, pandemic fatigue and personal experiences continue to shape perceptions about infectious diseases risk, and trust in government entities and associated public health surveillance and response.

To address this information gap and support preparedness efforts, we developed a survey to explore the knowledge, attitudes, and practices of community members in Victoria, Australia. We asked participants about their understanding of emerging respiratory diseases, their willingness to take part in public health surveillance activities such as HHTIs and the acceptability of various study elements. We additionally investigated participants’ comfort towards including their companion animals in HHTIs. Through this survey, we sought to inform development of a fit-for-purpose local HHTI protocol (adapted from the WHO *Unity Studies* template protocol) that balances public health information needs with community perspectives. Project outputs will inform pilot studies during seasonal epidemics with public health partners and identify subpopulations to engage in future survey iterations and engagement activities. Thus, our research aims to support rapid activation of effective and high-quality HHTIs during future infectious disease outbreaks in Australia.

## Methods

### Study population and eligibility

We conducted a cross-sectional, anonymous and voluntary convenience survey of persons living in Victoria, Australia. To be eligible to take part in the survey, participants were required to be at least 18 years of age, and self-report that they lived in a private home with at least one other person at the time of survey completion. Persons reporting living in a hotel or motel, hostel or shared accommodation (e.g., a dormitory), closed residential setting (e.g., a residential aged care facility, rehabilitation facility, army barracks) or with no fixed abode, were ineligible. No additional eligibility criteria were applied.

### Survey design

The full survey instrument is available in the Supplementary Material. As the survey was anonymous, we did not collect personal information such as name, contact details, or home address. Age-group and residential postcode were collected to verify eligibility.

The survey (approximately 20–30 minutes) prevented progression if eligibility criteria were not met or consent was not provided. The survey included questions on: individual and household demographics; health practices, beliefs and sources of health information relating to respiratory diseases; and the acceptability of a hypothetical 14-day HHTI for human participants. The hypothetical scenario proposed data collection on travel history, medical history, and daily symptom status; and sample collection including nose and/or throat swabs on Days 1, 4, 7, and 9; and blood samples on Days 1 and 14, from both participants and their household members. Individuals reporting the presence of companion animals in the household (defined as “a pet or any other domestic animal that you care for, interact with regularly in your home or on your property, and may have responsibility for”) were asked additional questions on: interactions with companion animals in the household; health practices and sources of information relating to companion animals; and acceptability of having data and specimens collected from their companion animal in a HHTI. If participants reported having more than one species of companion animal, they were asked to nominate the species of their ‘main’ companion animal with which they had the most interaction and respond accordingly in subsequent questions.

Questions were asked in various formats, including binary (Yes/No), multiple-choice, ‘select all that apply’, 5-point Likert scales and open-ended free text fields to detail reasoning. Questions other than free text were mandatory. Specific response options for Likert scales were tailored to the question (e.g., ranging from *‘Very uncomfortable’* to *‘Very comfortable’* or ‘*Very acceptable’* to *‘Very unacceptable’*. Definitions of key terms and brief, supporting explanations were provided at the beginning of survey sections.

At the conclusion of the survey, participants were able to provide any feedback, download an information and glossary resource about HHTIs and infectious disease surveillance, and enter their personal information (name and email only) into a separate, optional survey for a chance to win one of twenty $50 (AUD) gift vouchers. An independent team member who did not have access to the main survey data managed this database and randomly selected the winners.

### Survey dissemination

Study data were collected and managed using Research Electronic Data Capture (REDCap) electronic data capture tools hosted at the University of Melbourne,(15) following comprehensive pre-piloting and testing by our research team.

The survey was disseminated through several channels to attempt to capture a broad range of participants within the general Victorian population. Briefly, this included online distribution through social media campaigns (news articles led by the Peter Doherty Institute for Infection and Immunity and the Australian Partnership for Preparedness Research on Infectious Disease Emergencies, APPRISE), local community Facebook groups, and the posting of flyers in community settings (e.g., community hubs and libraries in metropolitan Melbourne) and veterinary clinics. Each promotion strategy was linked to a separate database to assess response rates.

Due to the exploratory and hypothesis-generating nature of the study, we did not specify a target sample size *a priori*. Instead, the survey was active for 20 weeks from 16^th^ February 2025 – 7^th^ July 2025. Data collection was ceased after 20 weeks when the response rate from online recruitment and flyers yielded no further participation for two consecutive weeks.

### Data management

All cleaning and analyses of quantitative data were conducted in R statistical software version 4.3.1(16)

### Analysis

In accordance with our pre-specified statistical analysis plan (Supplementary Material), responses from consenting individuals who provided all demographic data, and responded to at least one of our outcomes of interest, were included in the analysis dataset, regardless of survey final completion status. Our outcomes of interest were: 1) willingness to participate in HHTIs (hypothetical 14-day HHTI as outlined above); 2) acceptability of each HHTI component (i.e., epidemiological and clinical data, provision of respiratory and other specimens); 3) acceptable frequency of respiratory and blood specimen collection; 4) acceptability of HHTI components for companion animals; and, 5) acceptability of specimen collection methods for companion animals.

We summarised participant demographic data as frequency and percentage for categorical variables, and median and interquartile range (IQR) for continuous variables. We dichotomised geographical region of residence into the metropolitan area of Greater Melbourne or the regional area, using the Greater Capital City Statistical Area (GCCSA) as defined by the Australian Bureau of Statistics (ABS).(17) Where a postcode crossed GCCSA boundaries, it was assigned to the area containing >50% of the population. Summary statistics of participant demographic data were compared to 2021 Victorian population census data to assess representativeness.(18) The frequency and percentage of each response option (e.g., ‘*Very likely’*) was reported for each outcome. We used Clopper-Pearson exact binomial 95% confidence intervals (CI) to quantify uncertainty around estimates.(19) We did not report p-values or conduct multivariable analyses given the exploratory nature of the survey. No additional analyses were completed to handle missing data.

We conducted a primary exploratory subgroup analysis to determine if overall willingness to participate in a HHTI (outcome one) differed by demographic subgroups including gender, age, geographical area, language background, Medicare status (the Australian public health care system for which all Australian citizens, permanent residents, and some temporary visitors are eligible), level of education, and the presence of children and pets in the household. Subcategories with less than 10 participants were aggregated for this subgroup analysis, including: a) older age groups into “*60+ years*”; b) Medicare responses of *Unsure* or *No* into “*Unsure or ineligible”; and*, c) education responses of *Primary School* or *Secondary School* into “*Primary or secondary school*”. Given the focus of these analyses is on respiratory pathogens, we *post-hoc* excluded responses relating to companion animals that do not breathe air (e.g., fish) from the companion animal analyses. We conducted a second exploratory *post-hoc* subgroup analysis to determine if acceptability for companion animal components (outcome four) differed among those who nominated a cat or dog as their main companion animal.

### Ethics

This study was approved by the Human Ethics Committee of the University of Melbourne (Ethics ID: 2025-41424-65037-5). Ethics approval covered the collection and analysis of study data, including subgroup analysis by key demographics (excluding by First Nations status).

## Results

We received 282 survey responses from February to July 2025, of which 235 were included in the final analysis dataset (Supplementary Figure 1). In brief, eight partial respondents did not consent or meet eligibility criteria, 26 participants did not provide all required demographic data, and an additional 13 responses were excluded as they did not contribute towards any outcomes of interest.

Compared to the general Victorian population, a higher proportion of participants in our analysis dataset reported being female (63.8% in our survey sample compared to 50.8% in Victoria), aged under 60 years (94.5% vs 56.2%), tertiary-educated (80.8% vs 28.5%), of Aboriginal or both Aboriginal or Torres Strait Islander heritage (49.0% vs 1.01%), born in Australia (79.1% vs 65.0%) and only speaking English only at home (79.1% vs 67.2%) (Table 1). The median household size was 3 (IQR 2–4), with participants most frequently reporting living with their partner (81.3%), children (42.1%) and/or parents (30.6%). Further, the majority of participants (79.1%) reported having at least one companion animal in the household, with dogs (n=92, 49.6%) and cats (n=80, 43.0%) most frequently reported. Individual and household characteristics are detailed further in Table 1.

**Table 1.** Characteristics of participants and their households included in the final analysis dataset, compared to Victorian Census Data 2021(20)

| Individual characteristic | n<sub>survey</sub>(%) | n<sub>census</sub>(%) |
| --- | --- | --- |
| *Gender* |  |  |
| Male | 83 (35.3%) | 49.2% |
| Female | 150 (63.8%) | 50.8% |
| Non-binary | 2 (0.9%) | - |
| *Age-group (years)* |  |  |
| 18-19 | 10 (4.3%) | 2.2% |
| 20-24 |  | 6.3% |
| 25-29 | 78 (33.2%) | 7.3% |
| 30-39 | 78 (33.2%) | 15.2% |
| 40-49 | 39 (16.6%) | 13.0% |
| 50-59 | 17 (7.2%) | 12.2% |
| 60-64 | 5 (2.1%) | 5.6% |
| 65-69 | 5 (2.1%) | 4.9% |
| 70-79 | 3 (1.3%) | 7.5% |
| 80+ | 0 (0.0%) | 4.4% |
| *Born in Australia* |  |  |
| Yes | 186 (79.1%) | 65.0% |
| *Language spoken at home* |  |  |
| English only | 186 (79.1%) | 67.2% |
| *Eligible for Medicare* |  |  |
| Yes | 212 (90.2%) | - |
| No | 19 (8.1%) | - |
| Unsure | 4 (1.7%) | - |
| *Aboriginal or Torres Strait Islander status* |  |  |
| Not Aboriginal and/or Torres Strait Islander | 116 (49.4%) | 94.5% |
| Aboriginal | 102 (43.4%) | 0.96% |
| Torres Strait Islander | 7 (3.0%) | 0.03% |
| Both Aboriginal and Torres Strait Islander | 6 (2.6%) | 0.02% |
| Prefer not to say | 4 (1.7%) | 4.5%* |
| *Highest level of education* |  |  |
| Primary school or lesser | 2 (0.9%) | 3.2% |
| Secondary school | 9 (3.8%) | 35.9% |
| Vocational training | 10 (4.3%) | 24.1%^ |
| Certificate/Diploma/technical qualification | 24 (10.2%) |  |
| Bachelor's Degree | 130 (55.3%) | 29.2%^ |
| Postgraduate degree (e.g., Master's, PhD) | 60 (25.5%) |  |
| *Employment status* |  |  |
| Currently employed | 205 (87.2%) | 87.5%<sup>§</sup> |
| Not currently employed | 24 (10.2%) | 4.6%<sup>§</sup> |
| Prefer not to say | 6 (2.6%) | 7.9%<sup>§</sup> |
| **Household characteristic** | **n<sub>survey</sub> (%)** | **n<sub>census</sub> (%)** |
| *Household location* |  |  |
| Greater Melbourne<sup>1</sup> | 195 (83.0%) | 75.6% |
| *Household size* |  |  |
| Median (IQR) | 3 (2,4) |  |
| *Household composition* |  |  |
| Households with persons under 18 years of age | 81 (34.5%) |  |
| Households with persons over 65 years of age | 51 (21.7%) |  |
| At least one companion animal | 186 (79.1%) |  |
| *Main companion animal in household* |  |  |
| Dog | 92 (49.6%) |  |
| Cat | 80 (43.0%) |  |
| Birds (excluding poultry) | 3 (1.6%) |  |
| Poultry (e.g., backyard chickens, ducks) | 7 (3.8%) |  |
| Small mammals (e.g., rabbits, guinea pigs) | 2 (1.1%) |  |
| Other<sup>2</sup> | 2 (1.1%) |  |

Source: Australian Bureau of Statistics, TableBuilder (20, 21)
<sup>1</sup> As defined by the Greater Capital City Statistical Area
<sup>2</sup> Two participants reported 'goldfish' for their main companion animal. Responses are excluded from companion animal questions
- Data are not available in census data
\* Not stated in census data
^ Of responses from persons >15 years of age. Note secondary school includes any secondary school completion. Primary school or lesser includes 'no educational attainment' or level of education inadequately described.
§ % of people reporting being in the labour force for full-time or part-time work (>15 years of age, not retired)

Participants were generally positive about participating in a 14-day hypothetical HHTI (outcome 1, Figure 1). Most participants (62.9%) expressed that they would be ‘*Very likely’* (23.8%, 95% CI: 18.5–29.8%) or ‘*Somewhat likely’* (39.1%, 95% CI: 32.9–45.7%) to take part. In contrast, 16.2% of participants indicated they would be ‘*Very unlikely’* (9.4%, 95% CI: 6.0– 13.8%) or ‘*Somewhat unlikely’* (6.8%, 95% CI: 3.9–10.8%) to participate. The confidence intervals for each response category show an overall trend of willingness towards participation, rather than reluctance (Figure 1).

The exploratory subgroup analysis (Supplementary Table 1) highlighted one demographic subgroup that may be more in favour of HHTI participation. Most participants with children in the household (n=61, 75.3%) reported being ‘*Very likely*’ or ‘*Somewhat likely*’ to participate in a HHTI, compared to participants with no children in the household (n=87, 56.5%). No other notable differences were identified.

**Figure 1:**
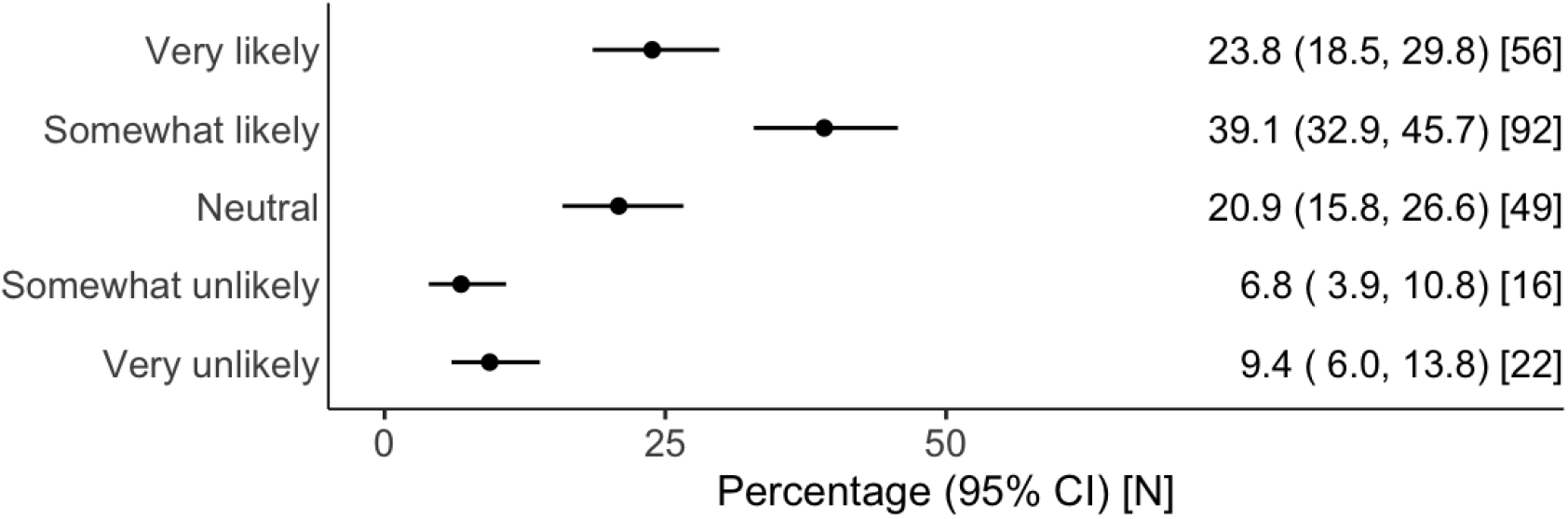
Participant willingness to take part in a specified hypothetical household transmission investigation (HHTI) scenario, including epidemiological and clinical data and sample (nasal and/or throat swabs and blood) collection over a 14 day period.

Most participants were *‘Very comfortable’* or *‘Somewhat comfortable’* with providing epidemiological and clinical data (range: 75.4–84%) (Figure 2). Nearly half of the participants reported being *‘Very comfortable’* with collection of travel history and symptom status (47.4% and 47.8% respectively) compared to medical history (34.9%). The majority of participants also reported being *‘Very comfortable’* or ‘Somewhat comfortable’ with all specimen types, with slight difference observed for nose or throat swabs, saliva, phlegm (range: 56.1–77.9%) compared to urine and blood (range: 56.1–59.3%) (Figure 2). However, fewer participants felt *‘Very comfortable’* overall with respiratory and other specimen collection (range: 21.6–31.6%), compared to questionnaire-based data collection.

**Figure 2:**
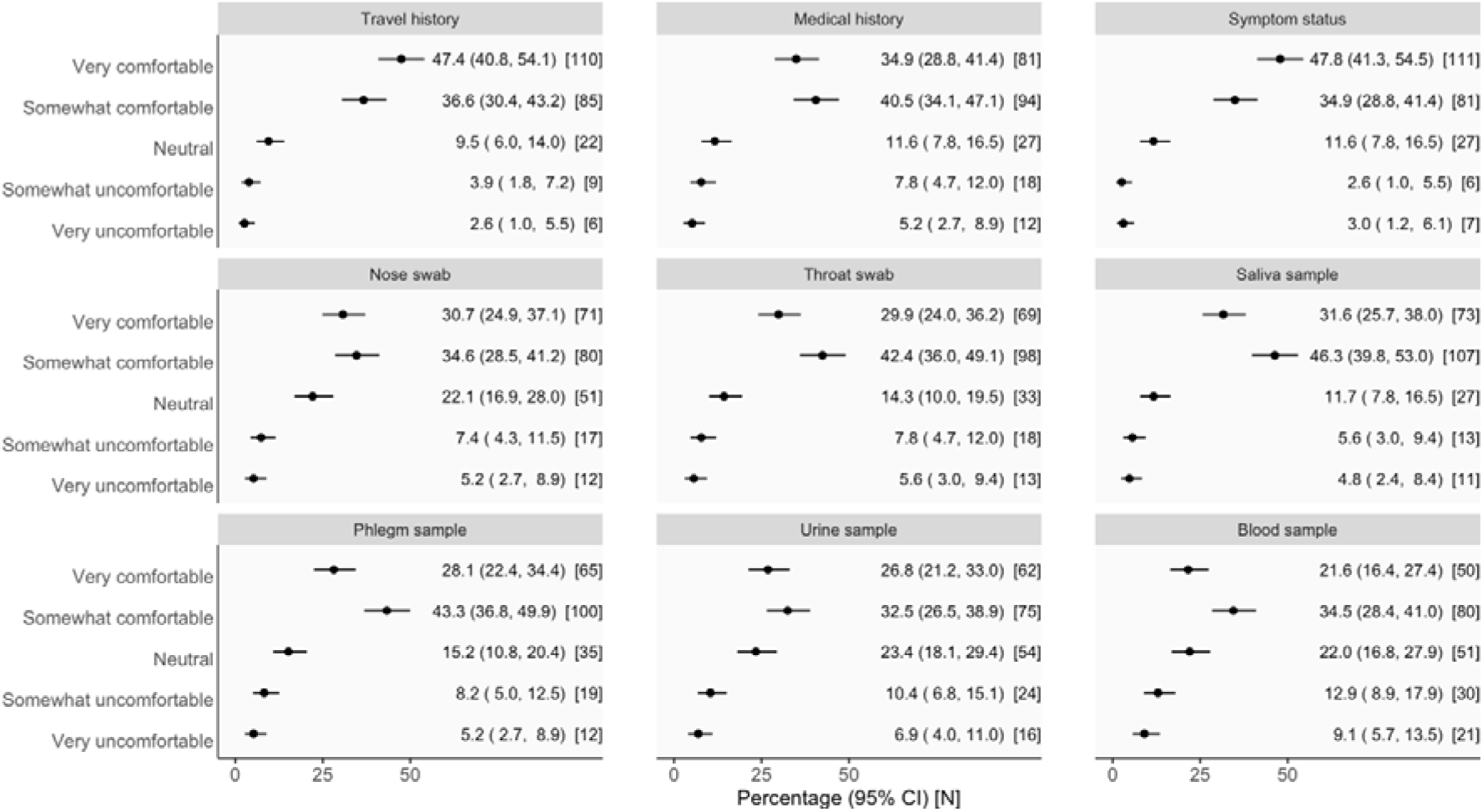
Participant comfort with components of household transmission investigations (HHTIs), including epidemiological and clinical data and sample (nasal and/or throat swabs and blood) collection.

Participants were notably more comfortable to provide more frequent nose and/or throat swabs compared to blood samples in a 14-day HHTI (Figure 3). The most favoured frequency of respiratory sample collection was two swabs, with just under 80% of participants indicating that they would be comfortable to provide two or more respiratory swabs in 14 days. Nearly one quarter indicated comfort with providing daily swabs. In contrast, most participants favoured providing only one blood sample in the 14 day period, with only 42.5% of participants comfortable with two or more samples.

**Figure 3:**
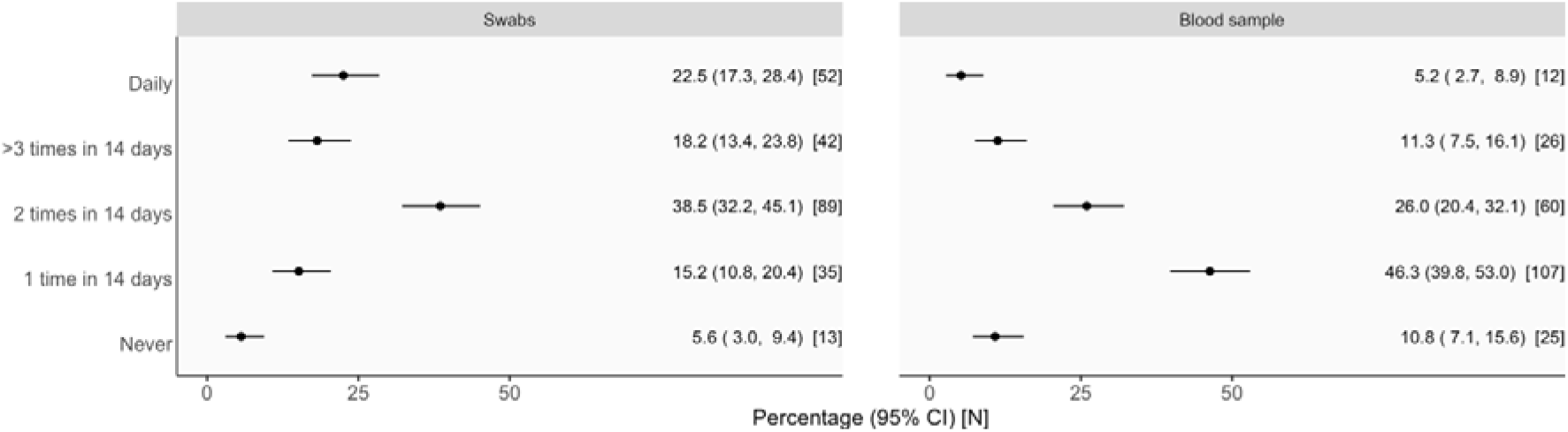
Participant preferences for frequency of respiratory swabs and blood sample collection from themselves in household transmission investigations (HHTIs).

Of the different respiratory specimen collection methods proposed, participants were most comfortable with collection of a self-swab for transport to a laboratory for testing via a courier (n=145, 62.8%), swabbing their own nose and/or throat for a rapid antigen test (RAT) (n= 134, 58.0%), or having a nurse or doctor visit their home for specimen collection (n= 114, 49.4%) (Supplementary Table 2).

The factors that would most motivate participants to take part in a HHTI included clear explanation of data usage (n=162, 70.4%), assurance of data privacy (n=157, 68.3%) and being paid for their time (n=146, n=63.5%) (Supplementary Table 3). Just under half (40.0%) of participants indicated that fewer tests or data collection points would motivate their participation, with several participants specifically noting in additional free text fields that they or other household members had a phobia of needles and/or acknowledged the complexity of blood sample collection from children.

Most participants (n=141, 61.3%) indicated that they or their household members would require a form of support to participate in a HHTI. This included support for self-testing and/or travelling to a clinic for testing (n=111, 48.3%), support to understand study requirements and communicate with the study team (n=77, 33.5%) or access to study instructions (n=75, 32.6%).

Extension of data and specimen collection to companion animals was viewed similarly to that in humans. Most participants reported they were either ‘*Very comfortable’* or *‘Somewhat comfortable’* with the collection of their companion animal’s medical history and signs of illness (more than 80%), followed by the collection of nose or throat swabs (around 70%) with lower, but still notable, comfort with more invasive blood samples (63.1%) being taken from their companion animals (Figure 4). Subgroup analysis showed that participants who nominated a dog as their main companion animal were more likely to report being *‘Very comfortable’* with each HHTI component compared to those who nominated a cat (7.3–11.9 percentage points higher), though overall comfort (‘*Very comfortable’* and *‘Somewhat comfortable’*) was similar (Supplementary Table 4).

**Figure 4:**
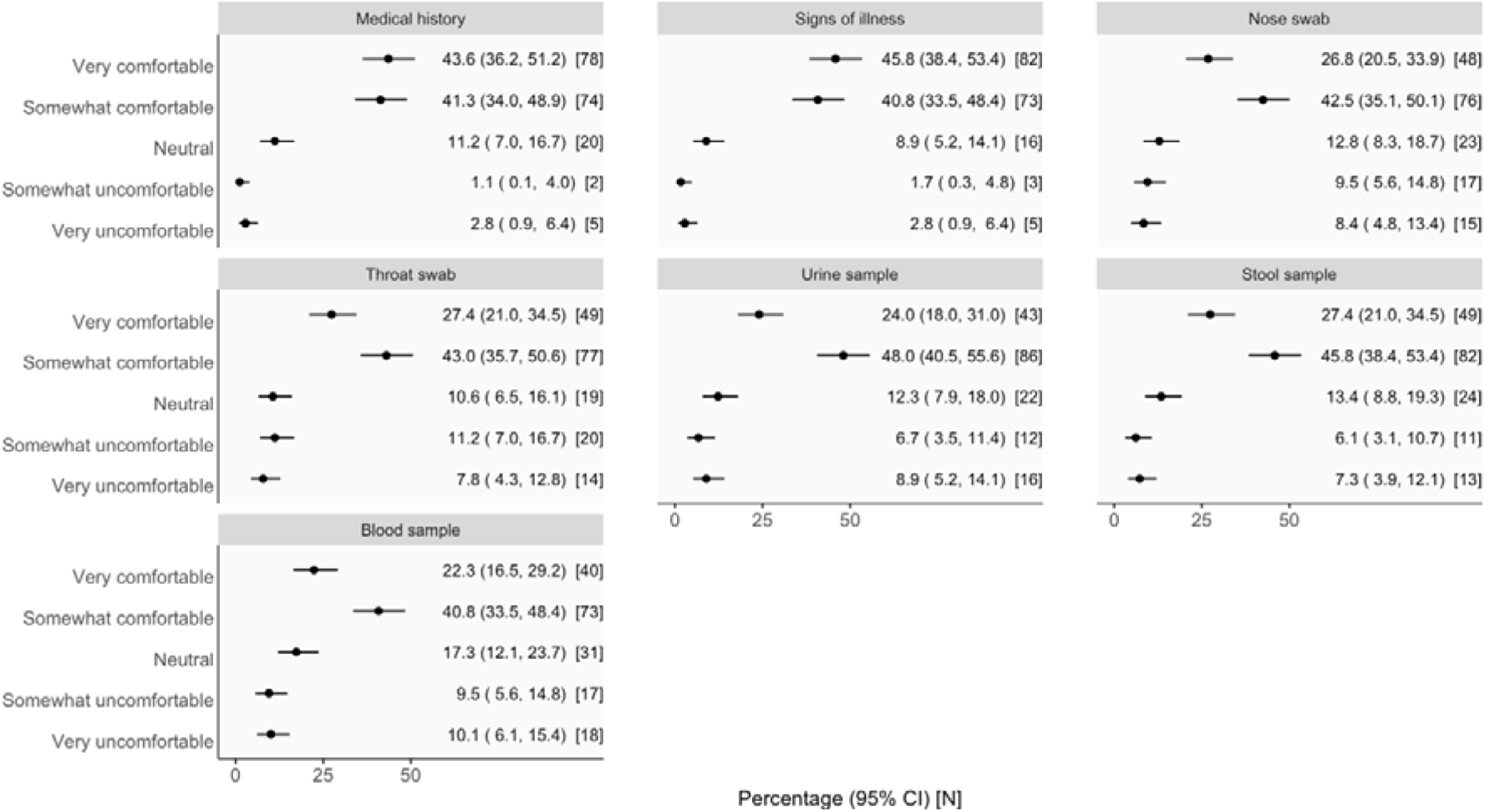
Participant comfort in providing epidemiological and clinical data, and specimens collected from their companion animals in household transmission investigations (HHTIs).

Acceptability for companion animal swabbing (*‘Very acceptable’* or *‘Somewhat acceptable’*) was very high (72.5–78.7%) and did not differ between owner- and veterinarian/veterinary nurse-collection, or between at-home and laboratory testing methods (Figure 5). There were no apparent differences in acceptability whether the main companion animal was a dog or a cat (Supplementary Table 5).

**Figure 5:**
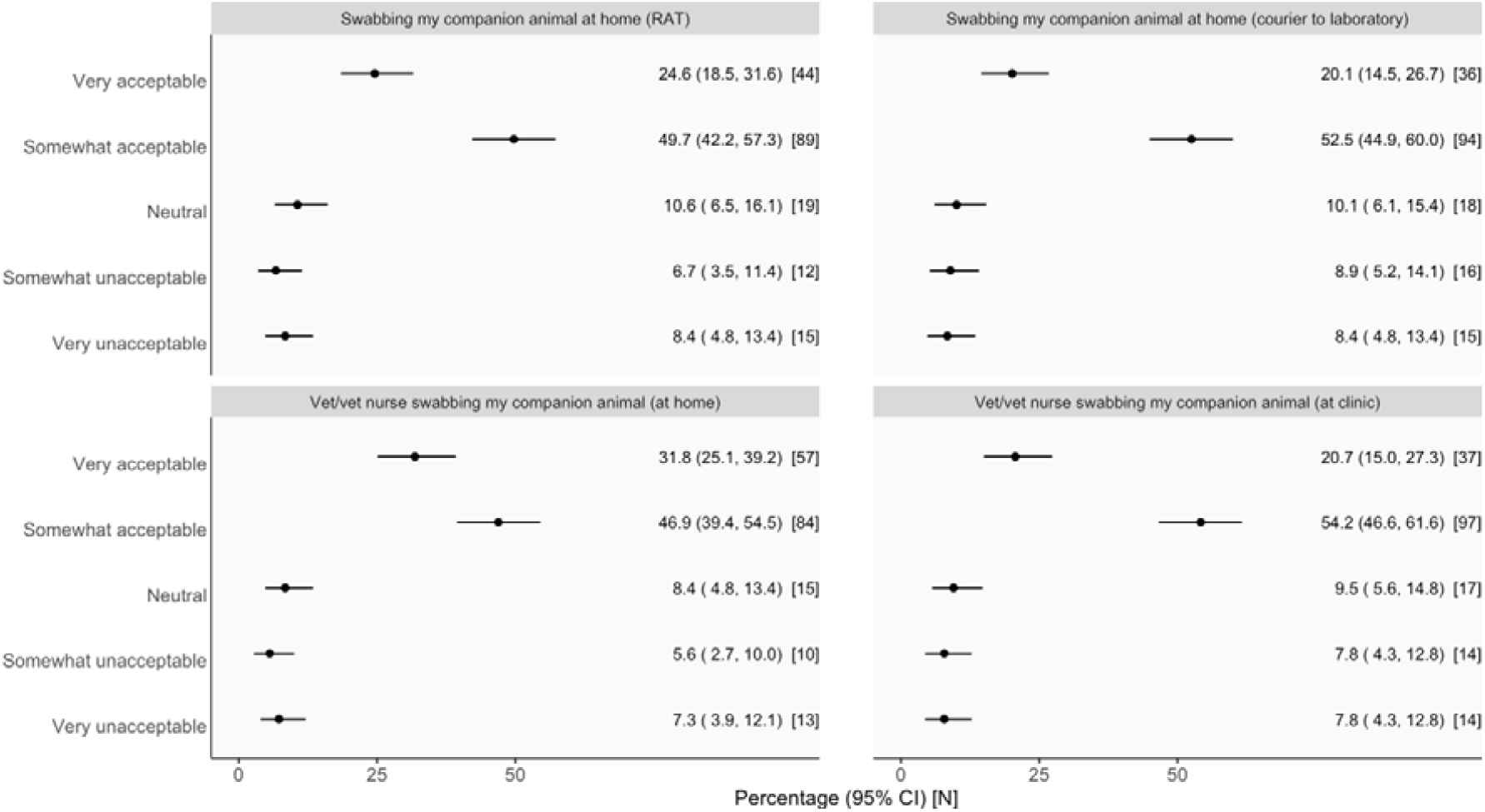
Participant preferences for method of respiratory specimen collection from their companion animals in household transmission investigations (HHTIs).

## Discussion

Our study generated novel data on the acceptability of HHTI participation among adults living in Victoria, Australia, from which to benchmark future national and international studies. Participants indicated high levels of comfort and acceptability towards our hypothetical 14-day HHTI involving epidemiological and clinical data collection and four nose/throat swabs for testing. We found differences in acceptability between different HHTI components — participants were most comfortable with the collection of clinical and epidemiological data, with lesser but still high overall comfort with providing multiple (two or more) respiratory (nasal, throat, or saliva) specimens in a 14-day period. Unsurprisingly, participants were least comfortable with potentially sensitive specimens such as urine and more invasive specimens such as blood samples. Our exploratory subgroup analysis demonstrated that willingness to participate was slightly increased among participants with children in their household, noting no statistical comparisons were made due to the exploratory nature of the survey. The reported acceptability of involvement of companion animals in HHTIs was notably as high as human-focused components.

Respondents were not representative of the broader Victorian population with a higher proportion of participants being tertiary educated, younger, female, or identifying as Aboriginal and/or Torres Strait Islander. This may reflect our recruitment strategy which utilised scientific social media posts associated with local research organisations (including APPRISE) or in local community groups. The APPRISE consortium have a long-standing history of participatory research with First Nations communities, which may explain how we achieved an exceptionally high response rate from this population group. While population representativeness has obvious advantages, overrepresentation of First Nations communities as we have achieved provides a unique opportunity for future *post-hoc* analyses. We may be able to generate robust insights on acceptability by First Nations status, provided analyses are conducted with appropriate community oversight and governance. We are developing collaborations with First Nations researchers and community members to determine interest and will seek relevant ethical approvals if this is deemed culturally appropriate and relevant. This research could be used to support development of adapted and culturally appropriate HHTIs and complement other engagement activities led by First Nations collaborators in other Australian jurisdictions.

Our recruitment strategy also included the physical distribution of flyers at a small number of veterinary clinics, which yielded modest participation, and community hubs, which yielded no responses. The contrast in response rates between online (social media) and in-person advertisements suggests that passive distribution methods in community settings may be less effective in engaging a representative subset of the population, especially those who are less digitally connected. Further, members of the Victorian population with diverse linguistic backgrounds, who may also be part of refugee or migrant communities (i.e., not born in Australia), may have been unable to participate as our recruitment materials and survey were only available in English. This potentially contributed further to our non-representative survey sample.

Population groups who experience inequitable infectious disease risks are typically under-represented in public health research and data. Future work should combine a wider online outreach strategy with more targeted use of in-person engagement approaches that ensures accessibility and ensure that diverse community perspectives are captured. This could involve offering translated materials and multiple formats for survey consent and completion (oral recorded vs. written). Active, relationship-based, and co-designed strategies are essential to ensure these methods of survey advertisement and dissemination are effective and culturally appropriate. Modifying our approach for future survey iterations will help capture richer data and support development of several culturally appropriate HHTI protocols. These protocols will support the collection of population-representative data for public health response and decision-making in infectious disease emergencies.

Trust, risk perception, and social norms shape willingness to comply with and engage in public health response measures including vaccination campaigns.(22-25) While community participation has been explored in participatory research contexts, few studies have examined proactive engagement in public health investigations of novel or re-emerging respiratory diseases themselves. Our findings extend this literature and provide a previously unavailable benchmark for community acceptability of proactive disease surveillance investigations, particularly HHTIs, that occur upstream to generate the high-quality evidence needed for proportionate public health responses. Willingness to contribute to HHTIs including provision of respiratory samples was high among our respondents. Given the recent COVID-19 pandemic, it is possible that participant familiarity with self-collection of respiratory samples and normalisation of self-administered RAT testing as a pre-requisite for social participation contributed to noted acceptability. It is possible that participant demographics, including higher levels of tertiary education and English language proficiency, may reflect higher health literacy and potentially explain our findings. Despite a clear pattern of overall willingness, almost two-thirds of participants indicated that participation in HHTIs would be enabled by provision of financial support. This suggests that willingness and health literacy alone may not translate directly into HHTI participation without adequate resourcing and consideration of participant needs.

The reported acceptability of companion animals in HHTIs was notably as high as human-focused components. This provides novel insights that directly support expansion of the traditional ‘household’ epidemiological unit to also include companion animals in studies of novel or re-emerging respiratory viruses. Integrated companion animal-human HHTIs of novel or re-emerging respiratory viruses, with either species as index cases, will strengthen community-based event surveillance at the animal-human interface.(26) Such investigations would support risk assessment through characterisation of bi-directional interactions and related inter-species respiratory disease transmission dynamics.(9) This is particularly important given continued emergence and global spread of zoonotic threats with pandemic potential (e.g., Nipah, Avian influenza) and the complex interfaces between human, animal and environmental health.

In future survey iterations with a larger, and more representative, survey sample, we may be able to make population-level inferences about the acceptability of HHTIs. Once available, this dataset could be analysed using appropriate statistical methods including multiple correspondence analysis to explore how participant responses cluster — particularly across related questions, including animal- and human-focused questions. Weighted regression analysis could also be used to generate population-level estimates of acceptability, comfortability and willingness (with associated measures of uncertainty) to inform protocol design and potential policy decisions about enhanced surveillance initiatives in future disease outbreaks.

Acceptability of HHTI study design, including for the necessary data and specimen collection, is key to the participation and retention of study participants, and for the success of the study. Improving our understanding of the limitations and constraints on these aspects of enhanced surveillance platforms will improve the design of such studies and ultimately enable them to generate high-quality data, leading to improved, rapid characterisation of novel pathogens upon their emergence. The design of HHTIs can be explored within these constraints *a priori* to determine how to best allocate limited resources (e.g., considering the number and timing of tests as well as preferences for type of test and testing methods)(27, 28) in real-world studies to establish testing schedules that are considered acceptable by the community and enabling appropriate characterisation of specific outcomes (e.g., the secondary attack rate).

## Conclusions

To our knowledge, this is the first study exploring community willingness and attitudes towards participation in public health surveillance and respiratory virus testing in Australia, and globally. Although exploratory, our findings provide useful insights into the acceptability of HHTIs in Victoria, Australia from which to benchmark necessary future surveys and activities both within Australia and globally. Survey findings will be directly integrated into local HHTI protocols and piloted in seasonal respiratory epidemics to gather further define implementation plans and seek further community-input on feasibility. This will improve the ability of HHTIs to generate meaningful data to inform public health responses in future infectious diseases emergencies.

## Data Availability

Study data may be requested for related research. Please contact the corresponding author to discuss data availability and alignment with ethics approvals.

## List of abbreviations

ABS: Australian Bureau of Statistics
CI: Confidence Interval
GCCSA: Greater Capital City Statistical Area
HHTI(s): Household Transmission Investigation(s)
IQR: Interquartile Range
RAT: Rapid Antigen Test
REDCap: Research Electronic Data Capture
PCR: Polymerase Chain Reaction
WHO: World Health Organization

## Declarations

### Ethics approval and consent to participate

This study was approved by the Human Ethics Committee of the University of Melbourne (Ethics ID: 2025-41424-65037-5). Participant consent was embedded within the survey, following an introduction to the purpose of the survey and a link to a plain language statement. In line with our approval, consent was obtained through the survey itself. Commencing the survey and completing any subsequent questions was considered as the participant providing consent to take part in this research.

### Consent for publication

All authors provided consent and approved the manuscript for submission and publication.

### Authors’ contributions

Conceptualization: AJM, JPVC, DH, NM. Methodology: All authors. Software: All authors. Validation: AJM, JPVC. Formal analysis: AJM, MH, JPVC. Investigation: All authors. Data Curation: AJM, NM, MH. Writing – original draft: AJM, JPVC, MH, DH, NM. Writing – Review and Editing: All authors. Visualization: AJM, MH, DJP. Supervision: AJM, JPVC, DJP, JMcV. Project administration: AJM, DH. Funding acquisition: AJM, JMcV, DJP

## Acknowledgements

We wish to thank Jennifer MacLachlan (WHO Collaborating Centre for Viral Hepatitis, Peter Doherty Institute for Infection and Immunity) for her assistance in mapping postcodes to ABS statistical areas and for independently and randomly selecting survey prize winners.

