## Supplementary Material for "Understanding community knowledge, attitudes and practices related to participation in household transmission investigations during infectious disease outbreaks"

***Supplementary Figure 1:*** *CONSORT diagram showing survey participants and reasons for exclusion from analysis dataset*

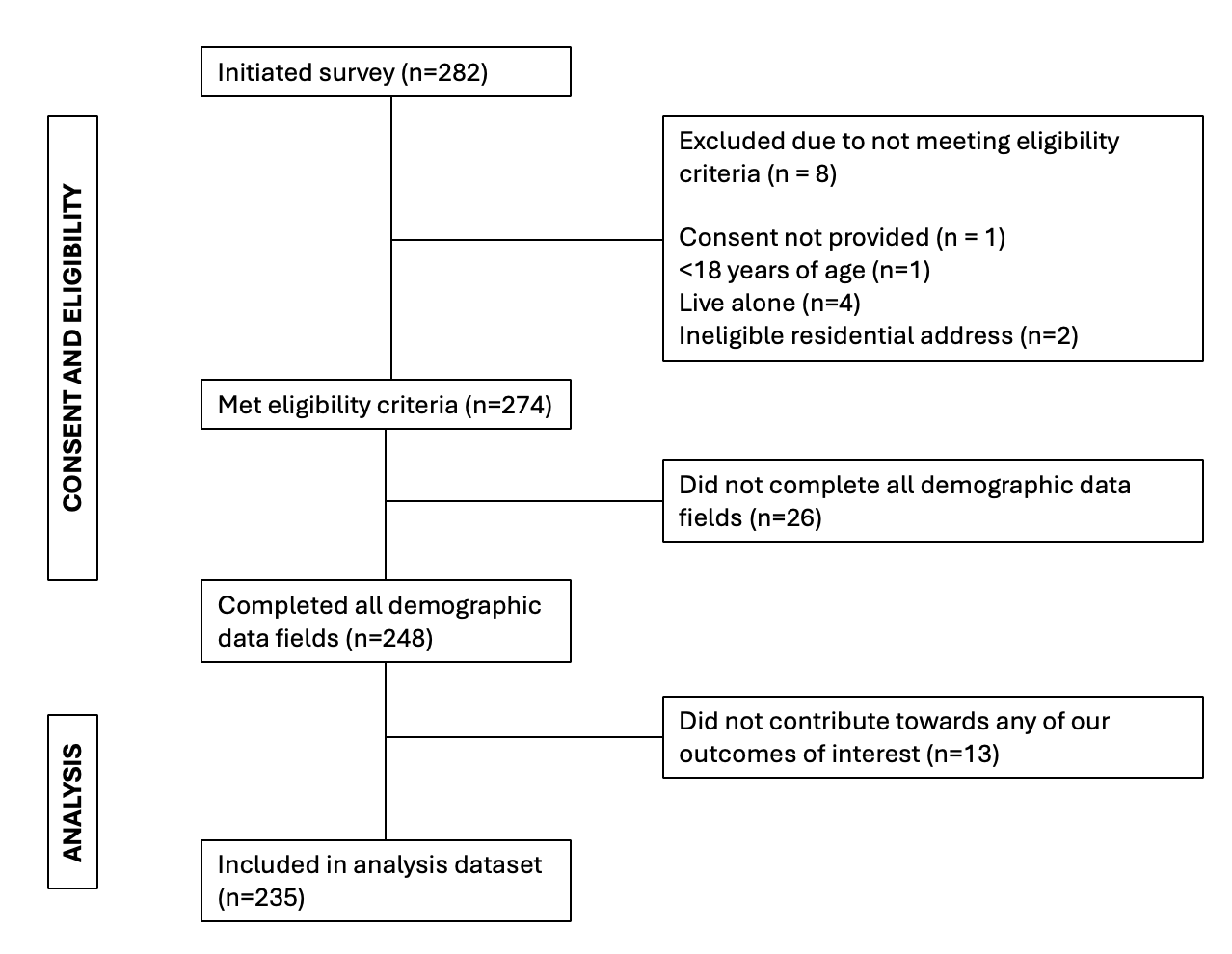

***Supplementary Table 1:*** *Subgroup analysis of overall participant willingness to take part in household transmission investigations (HHTIs). Participants were asked to respond in relation to a hypothetical HHTI design, which called for the following from them and their household members over a 14-day period: data collection including travel history, medical history, symptom status (daily); nose and/or throat swab collection on Days 1,4,7 and 9 of the study, and; blood sample collection on Days 1 and 14 of the study.*

| **Subgroup** | **Very likely (n, %)** | **Somewhat likely**  **(n, %)** | **Neutral**  **(n, %)** | **Somewhat unlikely**  **(n, %)** | **Very unlikely**  **(n, %)** |
| --- | --- | --- | --- | --- | --- |
| *Gender* | | | | | |
| Male | 14 (16.9%) | 42 (50.6%) | 19 (22.9%) | 4 (4.8%) | 4 (4.8%) |
| Female | 42 (28.0%) | 50 (33.0%) | 28 (19.0%) | 12 (8.0%) | 18 (12.0%) |
| *Age* | | | | | |
| 18-24 | 4 (40.0%) | 3 (30.0%) | 1 (10.0%) | 0 (0%) | 2 (20.0%) |
| 25-29 | 9 (11.5%) | 43 (55.1%) | 18 (23.1%) | 3 (3.8%) | 5 (6.4%) |
| 30-39 | 25 (32.1%) | 28 (35.9%) | 15 (19.2%) | 3 (3.8%) | 7 (9.0%) |
| 40-49 | 10 (26.0%) | 8 (21.0%) | 9 (23.0%) | 7 (18.0%) | 5 (13.0%) |
| 50-59 | 5 (29.4%) | 7 (41.2%) | 3 (17.6%) | 1 (5.9%) | 1 (5.9%) |
| 60+ | 3 (23.0%) | 3 (23.0%) | 3 (23.0%) | 2 (15.0%) | 2 (15.0%) |
| *Greater Capital City Statistical Area* | | | | | |
| Greater Melbourne | 42 (21.5%) | 76 (39.0%) | 42 (21.5%) | 14 (7.2%) | 21 (10.8%) |
| Rest of Victoria, Australia | 14 (35.0%) | 16 (40.0%) | 7 (17.5%) | 2 (5.0%) | 1 (2.5%) |
| *Languages spoken at home* | | | | | |
| English only | 45 (24.2%) | 71 (38.2%) | 39 (21.0%) | 13 (7.0%) | 18 (9.7%) |
| Language other than English | 11 (22.4%) | 21 (42.9%) | 10 (20.4%) | 3 (6.1%) | 4 (8.2%) |
| *Eligible for Medicare* | | | | | |
| Yes | 49 (23.1%) | 84 (39.6%) | 45 (21.2%) | 15 (7.1%) | 19 (9.0%) |
| No/Unsure | 7 (30.4%) | 8 (34.8%) | 4 (17.4%) | 1 (4.3%) | 3 (13.0%) |
| *Highest level of education* | | | | | |
| Primary or Secondary school | 1 (9.1%) | 4 (36.4%) | 3 (27.3%) | 2 (18.2%) | 1 (9.1%) |
| Vocational training | 2 (20.0%) | 3 (30.0%) | 4 (40.0%) | 1 (10.0%) | 0 (0%) |
| Certificate/Diploma/Technical qualification | 3 (12.5%) | 10 (41.7%) | 5 (20.8%) | 2 (8.3%) | 4 (16.7%) |
| Bachelor’s Degree | 26 (20.0%) | 56 (43.1%) | 31 (23.8%) | 7 (5.4%) | 10 (7.7%) |
| Postgraduate Degree (e.g., Master’s, PhD) | 24 (40.0%) | 19 (31.7%) | 6 (10.0%) | 4 (6.7%) | 7 (11.7%) |
| *Current employment status* | | | | | |
| Currently employed | 49 (23.9%) | 81 (39.5%) | 44 (21.5%) | 14 (6.8%) | 17 (8.3%) |
| Not currently employed | 6 (25.0%) | 8 (33.3%) | 4 (16.7%) | 2 (8.3%) | 4 (16.7%) |
| *Any children in household* | | | | | |
| No | 24 (15.6%) | 63 (40.9%) | 42 (27.3%) | 9 (5.8%) | 16 (10.4%) |
| Yes | 32 (39.5%) | 29 (35.8%) | 7 (8.6%) | 7 (8.6%) | 6 (7.4%) |
| *Any pets in household* | | | | | |
| No | 11 (22.4%) | 16 (32.7%) | 14 (28.6%) | 4 (8.2%) | 4 (8.2%) |
| *Yes* | 45 (24.2%) | 76 (40.9%) | 35 (18.8%) | 12 (6.5%) | 18 (9.7%) |

***Supplementary Table 2:*** *Participant comfort with respiratory specimen collection methods in HHTIs*

| ***Question*** | **n (%)**  **N=231** |
| --- | --- |
| *Which of the following methods for providing nose and/or throat swabs for respiratory diseases would you be comfortable with in a HHTI?* | |
| Swabbing your own nose and/or throat at home to do a rapid antigen test | 134 (58.0%) |
| Swabbing your own nose and/or throat at home and have it collected by a courier for testing | 145 (62.8%) |
| Having a nurse or doctor come to my home to swab my nose and/or throat for testing | 114 (49.4%) |
| Attending a health facility for a nurse or doctor to swab my nose and/or throat for testing | 64 (27.7%) |
| Attending a drive-through testing clinic for a nurse or doctor to swab my nose and/or throat for testing | 45 (19.5%) |
| None of the above | 9 (3.9%) |

***Supplementary Table 3:*** *Motivators for participating in HHTIs*

| ***Question*** | **n (%)**  **N=230** |
| --- | --- |
| *Which of the following factors would motivate you to take part in a HHTI?* | |
| Clear explanation of how my data will be used | 162 (70.4%) |
| Assurance of data privacy | 157 (68.3%) |
| Fewer tests or data collection points were required | 92 (40.0%) |
| Being paid for your time | 146 (63.5%) |
| Study being led by a research institute or university | 106 (46.1%) |
| Receiving up-to-date information about the disease from researchers who know the most about it | 93 (40.4%) |
| Receiving new or emerging treatments before the general population | 47 (20.4%) |
| Other | 9 (3.9%) |
| None of the above | 8 (3.5%) |

***Supplementary Table 4:*** *Subgroup analysis of participant comfortability with their nominated main companion animal (dog or cat) to take part in components of household transmission investigations (HHTIs).*

| **Component/**  **Main companion animal** | **Very**  **comfortable**  **(n, %)** | **Somewhat**  **comfortable**  **(n, %)** | **Neutral**  **(n, %)** | **Somewhat**  **uncomfortable**  **(n, %)** | **Very**  **uncomfortable**  **(n, %)** | **Missing** |
| --- | --- | --- | --- | --- | --- | --- |
| *Medical History* | | | | | |  |
| Cat | 32 (41.6%) | 37 (48.1%) | 6 (7.8%) | 0 (0%) | 2 (2.6%) | 3 |
| Dog | 44 (48.9%) | 31 (34.4%) | 11 (12.2%) | 2 (2.2%) | 2 (2.2%) | 2 |
| *Signs of illness* | | | | | | |
| Cat | 31 (40.3%) | 38 (49.4%) | 6 (7.8%) | 0 (0%) | 2 (2.6%) | 3 |
| Dog | 47 (52.2%) | 31 (34.4%) | 8 (8.9%) | 2 (2.2%) | 2 (2.2%) | 2 |
| *Nose swab* | | | | | | |
| Cat | 17 (22.1%) | 40 (51.9%) | 5 (6.5%) | 8 (10.4%) | 7 (9.1%) | 3 |
| Dog | 29 (32.2%) | 31 (34.4%) | 15 (16.7%) | 8 (8.9%) | 7 (7.8%) | 2 |
| *Throat swab* | | | | | | |
| Cat | 17 (22.1%) | 40 (51.9%) | 4 (5.2%) | 10 (13.0%) | 6 (7.8%) | 3 |
| Dog | 29 (32.2%) | 33 (36.7%) | 13 (14.4%) | 8 (8.9%) | 7 (7.8%) | 2 |
| *Urine sample* | | | | | | |
| Cat | 15 (19.5%) | 43 (55.8%) | 5 (6.5%) | 5 (6.5%) | 9 (11.7%) | 3 |
| Dog | 26 (28.9%) | 38 (42.2%) | 14 (15.6%) | 6 (6.7%) | 6 (6.7%) | 2 |
| Stool sample | | | | | | |
| Cat | 18 (23.4%) | 41 (53.2%) | 8 (10.4%) | 3 (3.9%) | 7 (9.1%) | 3 |
| Dog | 28 (31.1%) | 37 (41.1%) | 13 (14.4%) | 7 (7.8%) | 5 (5.6%) | 2 |
| Blood sample | | | | | | |
| Cat | 14 (18.2%) | 38 (49.4%) | 9 (11.7%) | 6 (7.8%) | 10 (13.0%) | 3 |
| Dog | 25 (27.8%) | 28 (31.1%) | 20 (22.2%) | 10 (11.1%) | 7 (7.8%) | 2 |

***Supplementary Table 5:*** *Subgroup analysis of participant acceptability for methods of respiratory specimen collection from their nominated main companion animal (dog or cat) within household transmission investigations (HHTIs).*

| **Collection method/**  **Main companion animal** | **Very**  **acceptable**  **(n, %)** | **Somewhat**  **acceptable**  **(n, %)** | **Neutral**  **(n, %)** | **Somewhat**  **unacceptable**  **(n, %)** | **Very**  **unacceptable**  **(n, %)** | **Missing** |
| --- | --- | --- | --- | --- | --- | --- |
| *Swabbing my companion animal at home (RAT)* | | | | | |  |
| Cat | 14 (18.2%) | 46 (59.7%) | 3 (3.9%) | 8 (10.4%) | 6 (7.8%) | 3 |
| Dog | 28 (31.1%) | 38 (42.2%) | 12 (13.3%) | 4 (4.4%) | 8 (8.9%) | 2 |
| *Swabbing my companion animal at home (courier to laboratory)* | | | | | | |
| Cat | 16 (20.8%) | 44 (57.1%) | 3 (3.9%) | 8 (10.4%) | 6 (7.8%) | 3 |
| Dog | 18 (20.0%) | 46 (51.1%) | 11 (12.2%) | 7 (7.8%) | 8 (8.9%) | 2 |
| *Vet/vet burse swabbing my companion animal (at home)* | | | | | | |
| Cat | 20 (26.0%) | 46 (59.7%) | 4 (5.2%) | 3 (3.9%) | 4 (5.2%) | 3 |
| Dog | 33 (36.7%) | 35 (38.9%) | 8 (8.9%) | 5 (5.6%) | 9 (10.0%) | 2 |
| *Vet/vet burse swabbing my companion animal (at clinic)* | | | | | | |
| Cat | 15 (19.5%) | 46 (59.7%) | 5 (6.5%) | 5 (6.5%) | 6 (7.8%) | 3 |
| Dog | 18 (20.0%) | 48 (53.3%) | 9 (10.0%) | 8 (8.9%) | 7 (7.8%) | 2 |

***Other supplementary material***

***Questionnaires/Survey instrument***

Thank you for your interest in this survey.

Researchers at the University of Melbourne are conducting a survey to understand your thoughts about diseases like the common cold and flu.

You can complete this survey on any computer or phone with a stable internet connection.

The required study information is below, and on the Plain Language Statement.

[link].

We want to know:

- how and where you access health information;
- whether you would be willing to take part in future studies to understand how respiratory diseases are and how they spread in households, and;
- if applicable, how you interact with any animals living on your property.

Your responses will inform how we work with our collaborators to learn about new respiratory diseases and support emergency public health responses.

This survey will take approximately 30 minutes to complete. By completing the entire survey, you will be eligible to enter a prize draw to win one of twenty $50 gift cards (see details below).

To take part in this survey you must:

- be at least 18 years of age;
- live in Victoria, Australia, and;
- currently live with at least one other person in a private home (e.g., house, apartment, unit or similar). For the purposes of this study, this means that someone else has spent more than 3 nights in the same home as you in the past week, and;
- read the plain language statement (link) and consent to take part.

The information you provide will only be used for the purposes of this study. You may submit the survey at any point, but only complete survey responses will be included in the prize draw.

Prize draw details:

If you wish to enter the prize draw, you may do so by entering your name and contact email in a separate form which you can access at the end of this survey. This information will be used only for distributing the gift cards. Any personal identifying details you provide for this purpose will not be linked to your survey responses in any way. They will be kept in a separate database and deleted once gift cards have been distributed.

Winners will be randomly selected from eligible participants.

Thank you for your time and participation.

Note: this study is approved by the University of Melbourne Human Research Ethics Committee (##31424) and is funded by a National Health and Medical Research Council grant.

Section 1: Individual consent and demographic details

1. Do you understand the purpose of the study as outlined above and in the Plain Language Statement? (Required)

- Yes
- No

2. Do you provide consent to take part in this survey? (Required)

- Yes
- *[IF YES],* Are you at least 18 years of age or older?
  - Yes
  - No
- No

3. Do you live alone or with other people? (Required)

- Alone
- With other people

4. What is the postcode of the suburb you live in?
5. What best describes your current living situation? (Required)

- Private home (house, apartment, unit or similar)
- Hotel or motel
- Hostel or shared accommodation outside of a private home (e.g., dormitory)
- Closed residential setting (e.g., aged care home, rehabilitation facility, army barracks).
- No fixed abode

6. What is your gender? (Required)
   - Male
   - Female
   - Non-binary
   - Other identity (please specify):

**TEXT BOX* (Internal: Not required)*

- Prefer not to say

7. How old are you? (Required)
   - 18-24
   - 25-29
   - 30-39
   - 40-49
   - 50-59
   - 60-64
   - 65-69
   - 70-79
   - 80+
8. Do you speak any languages other than English at home?

- Yes
- No [SKIP to Q10]

9. IF YES to Q8: Which of the following languages other than English do you speak at home? (Select all that apply)

- Arabic
- Cantonese
- Filipino / Tagalog
- French
- Greek
- Hindi
- Italian
- Mandarin
- Punjabi
- Sinhalese
- Spanish
- Vietnamese
- Other *TEXT BOX: “Please specify language(s). Separate each by a comma (,)*

10. Do you identify as Aboriginal or Torres Strait Islander? (Required)
    - No
    - Yes, Aboriginal
    - Yes, Torres Strait Islander
    - Yes, both Aboriginal and Torres Strait Islander
    - Prefer not to say
11. What best describes your migration status or background in Australia? (Required)
    - Born in Australia [SKIP TO Q15]
    - Arrived as a New Zealand citizen
    - Arrived as a skilled migrant
    - Arrived as a family migrant
    - Arrived as a refugee or through a humanitarian program
    - Arrived as an international student
    - Prefer not to say
    - Other (please specify):

**TEXT BOX**

12. What is your country of birth? (Required)

**DROP DOWN MENU**with *“My country of birth is not listed below”* and *“Prefer not to say”* at the top.

13. If “My country of birth is not listed below” is selected: Please specify your country of birth: *TEXT BOX*
14. In total, how long have you lived in Australia? (Required)

- Less than 1 year
- 1-5 years
- 6-10 years
- More than 10 years
- Prefer not to say

15. Are you currently eligible for Medicare? (Required)

- Yes
- No
- I am not sure
- Prefer not to say

16. What is the highest level of education/qualification you have completed? (Required)

- No formal education
- Primary school
- Secondary school
- Vocational training
- Certificate/Diploma/technical qualification
- Bachelor’s degree
- Postgraduate degree (e.g., Master’s, PhD)
- Other (please specify):

**TEXT BOX**

17. Are you currently employed?

- Currently employed [SKIP TO Q19]
- Not currently employed [GO TO Q18]
- Prefer not to say [SKIP TO Q20]

18. If “Not currently employed”: Do any of the following apply to you?

(Select all that apply)

- I am a student
- I am a stay-at-home parent/caregiver
- I am retired
- None of the above

19. What is your current occupation? (Required)

Select all that apply:

- Not currently employed/looking for work
- Studying
- Stay-at-home parent/caregiver
- Retired
- Manual worker or general labour role (e.g. cleaner, abattoir worker, etc.)
- Skilled tradesperson (e.g., plumber, electrician, carpenter, etc.)
- Service industry (e.g., retail, hospitality, security guard, etc.)
- Health care worker (e.g., nurse, GP, aged care worker, etc)
- Veterinarian or worker in agriculture sector (e.g. farmer, horticulturist, etc.)
- Specialist or technical role (e.g., engineering, IT, teacher, scientist, researcher, etc.)
  - Manager (e.g., supervisory or team management role, etc.)
  - Senior management or executive role (e.g., director, CEO, etc.)
  - Self-employed/business owner
  - Other (please specify):
    **TEXT BOX**

20. Have you ever been told by a doctor or a nurse that you have any of the following health conditions? (Required)

Select all that apply:

- Arthritis
- Asthma (not requiring medication)
- Severe asthma (requiring medication)
- Cancer (including remission)
- Dementia (including Alzheimer’s)
- Diabetes (excluding gestational diabetes)
- Heart disease (including heart attack or angina)
- Kidney disease
- Lung condition (including COPD or emphysema)
- Mental health condition (including depression or anxiety)
- Stroke
- Autoimmune diseases or disorders
- I have other long-term health condition(s), specify:
  **TEXT BOX**
- I have no long-term health conditions

Section 2: Household demographics and structure

Please keep the following definitions in mind when completing the rest of the survey:

A “household” is a group of people who live with you in the same home.

To “live with you” means that you have spent more than 3 nights in the same home in the past week.

21. INCLUDING yourself, how many people spent at least three nights in your home in the last week? For the purposes of this survey, this is the people that “live with you”. (Required)
22. Checkbox for 2, 3, 4, 5, 6, 7, 8, 9, 10, More than 10
23. Who lives with you in your home? (Required)

Select all that apply:

- Partner/spouse
- My child(ren)
- Parent(s)
- Grandparent(s)
- Sibling(s)
- Extended family (e.g., aunts, uncles, cousins)
- Friend(s)/housemate(s)
- Other (please specify): **TEXT BOX**

24. INCLUDING yourself, are there people aged 65 years or older in your home?

- Yes [GO TO Q24]
- No [SKIP TO Q25]

25. INCLUDING yourself, how many people who live with you are aged 65 years or older? (Required)
    **INTEGER**
26. Are any people who live with you under 18 years of age? (Required)
    - No [skip to Q26]
    - Yes

- If YES, How many people who live with you are pre-school aged children (under 5 years)?
  **INTEGER**
- If YES, How many people who live with you are primary school aged children (5-12)?
  **INTEGER**
- If YES, How many people who live with you are secondary school aged children (13-17)?
  **INTEGER**

27. Are there any pets (companion animals) or any other animals who live with you in your home or on your property? (Required)

Select all that apply:

- Cats
  - If SELECTED, how many cats?
    **INTEGER**
- Dogs
  - If SELECTED, how many dogs?
    **INTEGER**
- Birds excluding poultry (e.g., budgies)
  - If SELECTED, how many birds?
    **INTEGER**
- Poultry (e.g., backyard chickens and ducks)
  - If SELECTED, how many poultry?
    **INTEGER**
- Small pet mammals (e.g., rabbits, guinea pigs, ferrets, mice)
  - If SELECTED, how many small pet mammals?
    **INTEGER**
- Other (please specify the species and number):
  **TEXT BOX**
- None

Section 3: Attitudes and practices towards respiratory diseases, pandemics and health care seeking behaviour

The following questions relate to respiratory infectious diseases and your understanding of them. Please keep the following definitions in mind:

An “Infectious Disease” is an illness that is caused by germs (like bacteria or viruses) that can be spread from one person to another.

A “respiratory” infectious disease is an illness like the cold and flu, that affects your nose, throat and/or lungs. Symptoms from a respiratory disease may include a fever, cough, sore throat, runny nose, shortness of breath, headache, or loss of taste or smell. Examples of respiratory diseases include colds, flu, and COVID-19.

A “pandemic” is the global spread of a disease. Pandemics are often due to a new disease, like COVID-19, or a new strain of an existing disease, like Swine Flu in 2009. These can have a significant impact on human health causing large amounts of disease and death.

28. In general, how concerned are you about getting sick and having respiratory symptoms? (Required)

- Very concerned
- Somewhat concerned
- Neutral
- Not very concerned
- Not concerned at all

29. What are your main concerns about having respiratory symptoms? Please select up to three concerns. (Required)
    Select all that apply:

- Feeling unwell
- Passing it on to others
- Impact on routine, including social life & well-being
- Impact on work or education
- Financial costs due to loss of income or medical bills
- Impact on family and loved ones
- Being unable to access healthcare
- Long-term health effects
- None
- Other (please specify):
  **TEXT BOX**

30. How likely do you think another respiratory disease pandemic (like COVID-19) will occur in the next 10 years? (Required)

- Very likely
- Somewhat likely
- Neutral
- Somewhat unlikely
- Very unlikely

31. If there was another respiratory disease pandemic (like COVID-19), how concerned do you think would you be about the following? (Required)

| Scenario | Very concerned | Moderately concerned | Somewhat concerned | Slightly concerned | Not at all concerned |
| --- | --- | --- | --- | --- | --- |
| a. Your physical health | ⃝ | ⃝ | ⃝ | ⃝ | ⃝ |
| b. The physical health of other people in your home | ⃝ | ⃝ | ⃝ | ⃝ | ⃝ |
| c. Your well-being (e.g. social, financial, & mental health impacts) | ⃝ | ⃝ | ⃝ | ⃝ | ⃝ |
| d. The wellbeing (e.g. social, financial, & mental health impacts) of other people who live with you | ⃝ | ⃝ | ⃝ | ⃝ | ⃝ |

32. When you are sick or feeling unwell with respiratory symptoms, how often do you do the following? (Required)

| Behaviour | Always | Often | Sometimes | Rarely | Never |
| --- | --- | --- | --- | --- | --- |
| a. Seek health advice and information about your symptoms | ⃝ | ⃝ | ⃝ | ⃝ | ⃝ |
| b. Test yourself (e.g., using a RAT for COVID-19 or flu) | ⃝ | ⃝ | ⃝ | ⃝ | ⃝ |
| c. Stay away from other people in your home | ⃝ | ⃝ | ⃝ | ⃝ | ⃝ |
| d. Stay at home until you feel better | ⃝ | ⃝ | ⃝ | ⃝ | ⃝ |
| e. Take time off work | ⃝ | ⃝ | ⃝ | ⃝ | ⃝ |
| f. Wash your hands before preparing food for yourself or others | ⃝ | ⃝ | ⃝ | ⃝ | ⃝ |
| g. Wear a mask at home | ⃝ | ⃝ | ⃝ | ⃝ | ⃝ |
| h. Clean shared spaces at home (kitchen/bathroom) more frequently | ⃝ | ⃝ | ⃝ | ⃝ | ⃝ |
| i. Open windows and doors at home | ⃝ | ⃝ | ⃝ | ⃝ | ⃝ |
| j. Limit social gatherings with people outside the home | ⃝ | ⃝ | ⃝ | ⃝ | ⃝ |
| k. Avoid public transport | ⃝ | ⃝ | ⃝ | ⃝ | ⃝ |
| l. Wear a mask in public | ⃝ | ⃝ | ⃝ | ⃝ | ⃝ |

33. *[IF ALWAYS/OFTEN/SOMETIMES/RARELY TO A IN Q32]* When you seek health advice and information, what do you look for?? Select all that apply:

- Information about symptoms (how serious, how long they will last, etc)
- Information about when to see a doctor or nurse (including to get tested)What treatments are available
- How to protect others in your home from getting sick
- How to protect your community
- Other (please specify):

**TEXT BOX**

34. From which sources do you usually seek this information? (Required)
    Select all that apply:

- Doctor or nurse at your regular general practitioner / health clinic
- Urgent care clinic or walk in clinic
- Hospital emergency department
- Pharmacy and/or chemist
- Alternative medicine practitioners (e.g., homeopath, naturopath, traditional Chinese medicine practitioner, etc)
- Health hotlines (e.g., Nurse-on-Call)
- Friends or family
- Social media (e.g., Facebook, X (Twitter), Instagram, TikTok)
- Official websites (e.g., Department of Health, WHO, Better health network)
- Other websites through a search engine (e.g., Google, WebMD)
- Artificial Intelligence services (e.g., ChatGPT)
- Mainstream media (e.g., TV, radio)
- Friends and/or family
- Community/religious leader
- Other (please specify):

**TEXT BOX**

35. Why do you choose the above sources of information? (Required)
    **TEXT BOX** [SKIP TO Q37]
36. *[IF NEVER TO A IN Q32]* If you never usually look for information when you have cold or flu symptoms, could you please share your reasons why? (Required)
    **TEXT BOX**

Section 4: Animals in the household and interactions

[IF PERSON HAS INDICATED HAVING AT LEAST]

[For those who indicated in Q27 that they have just one companion animal species on their property, i.e., one of cats/dogs/birds/poultry/small companion mammals, show the following text and skip to Q39]

COMPANION ANIMALS

Please keep the following definitions in mind when completing the rest of the survey:

A “companion animal” refers to a pet or any other domestic animal that you care for and interact with regularly in your home or on your property, and may have responsibility for.

[For those who indicated in Q27 that they have more than one companion animal species on their property, show Q37]

For the remainder of this survey, please think about the companion animal that you previously indicated lives with you in your home or on your property.

Note: if you have more than one companion animal of the same species, please only think about the one that you have the closest interactions with for the rest of this survey.

37. You have selected that you have more than one species of pet or other animal on your property.

Please confirm the species of the animal that you consider to be your main companion animal (keeping in mind the definition above).

*Multiple choice options will appear here with participant’s responses to Q26*

E.g. 2 Cats, 1 Dog, Other: 1 Goldfish

38. [IF “Other” is selected in Q37]: Please confirm the species of your companion animal: *TEXT BOX*

For the remainder of this survey, please only think about the companion animal you selected above.

Note: if you have more than one companion animal of the same species, please only think about the one that you have the closest interactions with for the rest of this survey.

39. Do you think companion animals can get respiratory diseases from people? (Required)
    - Yes
    - No

Explain why you think this: **TEXT BOX**

40. Do you think people can get respiratory diseases from companion animals? (Required)
    - Yes
    - No

Explain why you think this: **TEXT BOX**

41. How often do you personally…? (Required)

|  | More than once a day | Once a day | More than once a week | Once a week | Less than once a week | Never |
| --- | --- | --- | --- | --- | --- | --- |
| a. Feed your companion animal | ⃝ | ⃝ | ⃝ | ⃝ | ⃝ | ⃝ |
| b. Clean up after your companion animal (e.g. picking up faeces) | ⃝ | ⃝ | ⃝ | ⃝ | ⃝ | ⃝ |
| c. Care for your companion animal (e.g. taking for exercise, giving medicine, cleaning their space) | ⃝ | ⃝ | ⃝ | ⃝ | ⃝ | ⃝ |
| d. Pat your companion animal | ⃝ | ⃝ | ⃝ | ⃝ | ⃝ | ⃝ |
| e. Cuddle your companion animal | ⃝ | ⃝ | ⃝ | ⃝ | ⃝ | ⃝ |
| f. Bring your companion animal onto your lap | ⃝ | ⃝ | ⃝ | ⃝ | ⃝ | ⃝ |
| g. Give kisses to your companion animal | ⃝ | ⃝ | ⃝ | ⃝ | ⃝ | ⃝ |
| h. Play with your companion animal with direct contact | ⃝ | ⃝ | ⃝ | ⃝ | ⃝ | ⃝ |

42. How often does your companion animal ...? (Required)

|  | More than once a day | Once a day | More than once a week | Once a week | Less than once a week | Never |
| --- | --- | --- | --- | --- | --- | --- |
| a. Lick your hands | ⃝ | ⃝ | ⃝ | ⃝ | ⃝ | ⃝ |
| b. Lick your face | ⃝ | ⃝ | ⃝ | ⃝ | ⃝ | ⃝ |
| c. Lick other parts of your body (e.g., arms/legs) | ⃝ | ⃝ | ⃝ | ⃝ | ⃝ | ⃝ |
| d. Bite or peck you | ⃝ | ⃝ | ⃝ | ⃝ | ⃝ | ⃝ |
| e. Scratch you | ⃝ | ⃝ | ⃝ | ⃝ | ⃝ | ⃝ |
| f. Share the same space with you (e.g., room, couch, bed, etc.) | ⃝ | ⃝ | ⃝ | ⃝ | ⃝ | ⃝ |

43. Do you and your companion animal interact in ways that are not listed above?

- Yes
- No

44. In what other ways do you and your companion animal interact? **TEXT BOX**
45. Does your companion animal live indoors (live with you inside your home) or outdoors (on your property)? (Required)

- Indoors only
- Outdoors only [skip to 48]
- Both indoors and outdoors

46. How much time does your companion animal usually spend outside (on your property)? (Required)

- My companion animal never goes outside
- My companion animal spends little time outside (e.g., to use the toilet or eat)
- My companion animal spends equal time inside and outside
- My companion animal spends most of their time outside

47. Where does your companion animal typically sleep at night? (Required)
    - In my or another household member’s bedroom (not on the bed)
    - In my or another household member’s bedroom (on the bed)
    - In a separate area of the home (e.g., laundry, living room)
    - Outside the home (e.g., kennel in backyard, on the porch/patio)
48. Does your companion animal go outside the boundaries of your property for walks or exercise? (Required)

- Yes
- No [skip to Q50]

49. Where does your companion animal go for walks or exercise? (Required). Select all that apply:

- Local streets/neighbourhood
- Local parks (including dog parks)
- Nature reserve/state parks
- Rivers or lakes
- Beaches
- Other (please specify):

**TEXT BOX**

50. Is your pet ever allowed to roam outside of the boundaries of your property by themselves? (Required)
    - Yes
    - No

51. In the last 12 months, have you seen your pet interact with wildlife and/or other non-domesticated animals or their carcasses? (Required)
    - No
    - Yes
      - If YES, Which species?
        - Birds
        - Mice/rats
        - Possums
        - Bats
        - Reptiles e.g., lizards
        - Other, please specify: **TEXT BOX**

- IF YES, How often do these interactions usually occur?
  - Daily
  - Weekly
  - Monthly
  - Less than once a month

Section 5: Companion animal health and information sources

52. What signs would make you think that your companion animal might be unwell with an infectious disease? (Required)
    Select all that apply:

- Changes in appetite and thirst
- Changes in behaviour and/or mood
- Changes in activity levels
- Respiratory symptoms (e.g., sneezing, coughing)
- Physical symptoms (e.g., limping, itching, rash, changes in coat, etc.)
- Diarrhoea or vomiting
- Changes in urination
- Discharge from eyes and/or nose
- Other (please specify): **TEXT BOX**
- None

53. When your companion animal begins to show signs of mild illness, what do you usually do first? (Required)

- Nothing
- Monitor for worsening symptoms before seeking advice
- Seek advice from trusted sources before booking a veterinary appointment
- Schedule a veterinary appointment
- Take them to an emergency veterinary service
- Other (please specify):

**TEXT BOX**

54. When your companion animal begins to show signs of severe illness, what do you usually do first? (Required)

- Nothing
- Monitor for worsening symptoms before seeking advice
- Seek advice from trusted sources before booking a veterinary appointment
- Schedule a veterinary appointment
- Take them to an emergency veterinary service
- Other (please specify): **TEXT BOX**

55. Have you taken your companion animal to the vet in the last 12 months?

- Yes
  - If YES, did the vet take a sample from your companion animal (for any reason)?
    - Yes
      - If YES, Which specimens were taken from your companion animal? (Select all that apply)
        - Ear swab
        - Nose swab
        - Throat swab
        - Blood sample
        - Rectal or faecal/stool sample
        - Skin sample (including lumps)
        - Urine sample
        - Other, please specify: **TEXT BOX**
    - No
- No

56. Do your interactions with your companion animal change when you are unwell?

- Yes
  - Please explain how your interactions changed: **TEXT BOX**
- No

57. Do your interactions with your companion animal change when your they are unwell?

- Yes
  - Please explain how your interactions changed: **TEXT BOX**
- No

Section 6: Attitudes Towards Household Transmission Investigations

The following questions relate to how you feel about household transmission investigations (referred to as HHTIs)

This is a type of study that collects health information from people who have respiratory diseases, and every person that lives in the same household as them. HHTIs provide information about how diseases spread, how severe they are, and what might be effective in preventing them.

These studies may collect the following information from you and people you live with over a two-week (14 day) period:

- Your travel history
- Your symptom status
- Your medical history
- A few swabs, blood and/or other samples for testing (see example below)

Example Testing Schedule:

- Nose/Throat swabs: collected on Days 1, 4, 7 and 9 of the study
- Blood samples: collected on Days 1 and 14 of the study

58. Based only on the information above, if you were asked today to take part in a HHTI, how likely would you be to do so? (Required)

- Very likely
- Somewhat likely
- Neutral
- Somewhat unlikely
- Very unlikely

59. Which of the following would you be comfortable to provide in a household transmission investigation? (Required)

| Information Type | Very comfortable | Somewhat comfortable | Neutral | Somewhat uncomfortable | Very uncomfortable |
| --- | --- | --- | --- | --- | --- |
| a. Your travel history | ⃝ | ⃝ | ⃝ | ⃝ | ⃝ |
| b. Your symptom status | ⃝ | ⃝ | ⃝ | ⃝ | ⃝ |
| c. Your medical history | ⃝ | ⃝ | ⃝ | ⃝ | ⃝ |
| d. Nose swab(s) | ⃝ | ⃝ | ⃝ | ⃝ | ⃝ |
| e. Throat swab(s) | ⃝ | ⃝ | ⃝ | ⃝ | ⃝ |
| f. Saliva sample(s) | ⃝ | ⃝ | ⃝ | ⃝ | ⃝ |
| g. Phlegm sample(s) | ⃝ | ⃝ | ⃝ | ⃝ | ⃝ |
| h. Blood sample(s) | ⃝ | ⃝ | ⃝ | ⃝ | ⃝ |
| i. Urine and/or other sample(s) | ⃝ | ⃝ | ⃝ | ⃝ | ⃝ |

60. Which of the following methods for providing nose and/or throat swabs for respiratory diseases would you be comfortable with in a HHTI? (Required)

Select all that apply:

- Swabbing your own nose and/or throat at home to do a rapid antigen test
- Swabbing your own nose and/or throat at home and have it collected by a courier for testing at a laboratory
- Having a nurse or doctor come to my home to swab my nose and/or throat for testing
- Attending a health facility for a nurse or doctor to swab my nose and/or throat for testing
- Attending a drive-through testing clinic for a nurse or doctor to swab my nose and/or throat for testing
- None of the above

61. How many times would you be comfortable providing the following samples during a household transmission investigation? For the purposes of this survey, this is how many times would you provide them during a 14-day study. (Required)

| Specimen Type | Daily | More than three times | Two times (i.e. once per week) | Only once | Never | Other (please specify) |
| --- | --- | --- | --- | --- | --- | --- |
| a. Nose and/or Throat swab(s) | ⃝ | ⃝ | ⃝ | ⃝ | ⃝ | *TEXT BOX* |
| b. Blood sample(s) | ⃝ | ⃝ | ⃝ | ⃝ | ⃝ | *TEXT BOX* |

62. Which of the following factors would motivate you to take part in a HHTI? (Required) Select all that apply:

- Clear explanation of how my data will be used
- Assurance of data privacy
- Fewer tests or data collection points were required
- Being paid for your time
- Study being led by a research institute or university)
- Receiving up-to-date information about the disease from researchers who know the most about it
- Receiving new or emerging treatments before the general population
- Other (please specify): **TEXT BOX**

63. Including you, would you or someone in your household require any of the following supports to participate in a HHTI? (Required)Select all that apply:

- None required.
- Support with self-testing and/or travelling to get tested at a clinic
- Support with communication (e.g., language interpreter, visual aids) and understanding the study requirements
- Support with accessing, or completing instructions
- Yes, other (please specify): **TEXT BOX**

[IF PREVIOUSLY INDICATED THAT THE HOUSEHOLD HAS ANIMALS ON THEIR PROPERTY]

64. Which of the following would you be comfortable to report or have collected in relation to your companion animal in a household transmission investigation? (Required)

| Information Type | Very comfortable | Somewhat comfortable | Neutral | Somewhat uncomfortable | Very uncomfortable |
| --- | --- | --- | --- | --- | --- |
| a. Signs of illness shown by my companion animal | ⃝ | ⃝ | ⃝ | ⃝ | ⃝ |
| b. Medical history | ⃝ | ⃝ | ⃝ | ⃝ | ⃝ |
| c. Nose swab(s) | ⃝ | ⃝ | ⃝ | ⃝ | ⃝ |
| d. Throat swab(s) | ⃝ | ⃝ | ⃝ | ⃝ | ⃝ |
| e. Blood sample(s) | ⃝ | ⃝ | ⃝ | ⃝ | ⃝ |
| f. Faecal/stool sample(s) | ⃝ | ⃝ | ⃝ | ⃝ | ⃝ |
| g. Urine sample(s) | ⃝ | ⃝ | ⃝ | ⃝ | ⃝ |

65. How acceptable are the following methods for the collection of nose and/or throat swabs from your companion animal in a HHTI ? (Required)

| Method of collection | Very acceptable | Somewhat acceptable | Neutral | Somewhat unacceptable | Very unacceptable |
| --- | --- | --- | --- | --- | --- |
| Swab my companion animal’s nose and/or throat at home to do a rapid antigen test | ⃝ | ⃝ | ⃝ | ⃝ | ⃝ |
| Swab my companion animal’s nose and/or throat at home to have it collected by a courier for testing at a laboratory | ⃝ | ⃝ | ⃝ | ⃝ | ⃝ |
| Have a vet/vet nurse come to my house to swab my companion animal’s nose and/or throat for testing (at no cost) | ⃝ | ⃝ | ⃝ | ⃝ | ⃝ |
| attending a vet clinic for a vet/vet nurse to swab my companion animal’s nose and/or throat for testing (at no cost) | ⃝ | ⃝ | ⃝ | ⃝ | ⃝ |

66. If you have any additional comments or questions about the contents of this survey, please share your thoughts below:

Section 7: End of survey

Thank you. We really appreciate the time you have taken to complete the survey.

If you would like to learn more about household transmission investigations, why they are important, and what is involved in them, you can download a resource sheet by clicking the link below:

[Link to HHTI Resource sheet]

If you wish to enter the draw to receive one of twenty $50 gift card, please submit your response and click on the link below:

[Link to prize draw entry form]

Please feel free to contact us at if you have any other questions or concerns regarding this survey.

***Statistical Analysis Plan***

This Knowledge, Attitudes and Practices (KAP) survey aims to understand what people in the community think about common respiratory diseases like the cold and flu, and how they seek healthcare when experiencing mild respiratory illness. The survey also asks several questions relating to broad study design of Household Transmission Investigations (HHTIs) protocols and willingness to participate (including with their companion animals). Insights will be used to help define adapted HHTI protocols that may be considered acceptable and feasible by the Victorian community.

The study objectives (below) are both technical and operational, to inform potential HHTI protocol adaptation as well as identify how to improve on our survey methodology in future related research projects.

The survey was disseminated through several channels to capture a broad range of participants and attempts to have a sample that is semi-representative of the general Victorian population. Briefly, the study was advertised on social media (APPRISE, Doherty communications), through community groups (online), vet clinics, and through community organisations (both online and in-person).

### Section 1: Objectives and relevant data

#### Primary objectives

1. Describe the demographics and characteristics of the population who engaged with our online survey.

Rationale: Will help us understand the representativeness of our survey sample and subsequent potential generalisability of findings by comparing to 2021 Victorian census data (Australian Bureau of Statistics. (2022). 2021 Census of Population and Housing, Victoria. Retrieved [7 July 2025] from <https://www.abs.gov.au/census/find-census-data/quickstats/2021/2>

.

2. Quantify willingness to participate in HHTIs (overall and by demographic subgroups).

Rationale: Will help us identify potential differences in responses by key demographic variables

3. Describe which components of HHTIs related to human participants (types of data or specimens, collection method/location and frequency of collection) may be considered acceptable by members of the Victorian community (overall and by demographic subgroups).

Rationale: Will provide insight into what people might be willing to do relating to their own participation in a HHTI and how frequently they are willing to do it.

4. Describe which components of HHTIs related to companion animal participants (types of data or specimens and collection method/location) may be considered acceptable by members of the Victorian community (overall and by demographic subgroups).

Rationale: Will provide insight into what people might be willing to do relating to their companion animals’ participation in a HHTI and how frequently they are willing to do it.

5. Describe reported ‘routine’ human-companion animal and companion animal-wildlife contact patterns such as type of and frequency of contact (overall and by demographic subgroups).

Rationale: Identify the types of interactions by pet type, to inform potential contact diary studies and/or further work

#### Secondary objectives

6. Assess survey completion rate (% completion once commenced) and time taken, and whether this differs by demographic subgroups. Explore where people stop in the survey.

Rationale: Will help us identify potential differences in completion rates by key demographic variables — is the survey itself accessible? Is there a particular point where the survey becomes disengaging?

7. Describe participants’ perceptions about future pandemics (overall and by demographic subgroups)

Rationale: Identify the knowledge and perceptions of our cohort to help inform future education and information campaigns.

8. Describe participants’ perceptions about zoonoses (overall and by demographic subgroups)

Rationale: Identify the knowledge and perceptions of our cohort to help inform future education and information campaigns.

9. Describe health care seeking behaviours and sources of trusted health information (overall and by demographic subgroups)

Rationale: In addition to understanding behaviours, may help us identify future channels for survey distribution and/or recruitment methods for HHTIs.

10. Describe health care seeking behaviours for companion animals (overall and by demographic subgroups)

Rationale: In addition to understanding behaviours, may help us identify how to extend HHTIs to include companion animals and what supports will be needed from veterinary partners.

11. Describe potential motivators for participation in HHTIs and supports required (overall and by demographic subgroups)

Rationale: Understand how to encourage participation and minimise potential impact associated with participating.

### Section 2: Outcome measures

Where feasible, responses relevant to each primary and secondary outcome measures will be reported overall and by primary subgroups of interest (as described in Data cleaning and analysis).

#### Primary outcome measures

1. Describe the demographics and characteristics of the population who engaged with our online survey.

   Table 1 contains all variables to be described for the survey population. In addition, the number of respondents from each survey source, and the number of people who initiated the survey and did not provide consent, will also be reported. Demographics will be compared to Victorian census data^1^ where available to understand potential representativeness of our participants.

Table 1. Demographic variables captured in the Household Transmission Investigation (HHTI) Knowledge, Attitudes and Practice (KAP) survey.

| Field | Response options | Complete^*^  n (%) | Incomplete^**^  n (%) | Overall  N | Redcap Q. Num (for ref) |
| --- | --- | --- | --- | --- | --- |
| Location | Metropolitan Melbourne vs rural/regional Victoria |  |  |  | 12 |
| Gender | Male, female, non-binary, prefer not to say |  |  |  | 18 |
| Age | 18 – 29, 30 – 49, 50 – 64,  ≥65 |  |  |  | 20 |
| Languages other than English spoken at home | No, Yes |  |  |  | 21 |
|  | If yes, language |  |  |  | 22, 23 |
| Aboriginal and Torres Strait Islander status | No, Yes - Aboriginal, Yes – Torres Strait Islander, Yes – both Aboriginal and Torres Strait Islander) |  |  |  | 24 |
| Migration status/background in Australia | Born in Australia, Arrived as a New Zealand citizen, Arrived as a skilled migrant, Arrived as a family migrant, Arrived as a refugee or through a humanitarian program, Arrived as an international student, Arrived for short term work, Prefer not to say, Other |  |  |  | 25,26 |
| Country of birth | Presented by WHO region |  |  |  | 27, 28 |
| Medicare status | Yes, No, I am not sure, Prefer not to say |  |  |  | 30 |
| Education | Primary school, secondary school, vocational training, certificate/diploma/technical qualification, Bachelor’s degree, Post-graduate degree (e.g. Master’s, PhD) |  |  |  | 31, 32 |
| Employment status^***^ | Currently employed, Not currently employed, Prefer not to say |  |  |  | 33  34, 35 |
| Household size | 2-10, More than 10 |  |  |  | 42 |
| Household composition^****^ | With and without kids (No, Yes)  If yes, median number of kids  With and without extended family and/or grandparents (No, Yes)  Friends/housemates (No, Yes) |  |  |  | 43, 44,  50-53  43, 44  43 |
| Pets or not | No, Yes  If yes, number/type reported |  |  |  | 56  59-64 |

* Participant consented and completed all mandatory fields
** Participant consented but did not complete all mandatory fields
*** Additional details available to report retirement/specific occupations if desired
**** Additional details available to report the number of individuals in household aged >65, <5, 5-12, 13-17 if desired

2. Quantify willingness to participate in HHTIs (overall and by demographic subgroups).

   Overall summary of willingness will be described by the question, “Based only on the information above, if you were asked today to take part in a HHTI, how likely would you be to do so?”, using a 5-point Likert scale ranging from “Very likely” to “Very unlikely”.
3. Describe which components of HHTIs related to human participants may be considered acceptable by members of the Victorian community (overall and by demographic subgroups).

   Survey respondents reported their willingness to provide data and specimens as part of their hypothetical involvement in a HHTI. This will be described by the question, “Which of the following would you be comfortable to provide about yourself in a household transmission investigation (HHTI)?”, using a 5-point Likert scale ranging from “Very comfortable” to “Very uncomfortable”.

Survey respondents reported how frequently they were willing to provide swabs or blood samples as part of their hypothetical involvement in a HHTI. This will be described by the question, “How many times would you be comfortable providing the following samples during a household transmission investigation?”, using Likert scale options “Daily, More than three times, two times (once per week), only once, never, other”.

4. Describe which components of HHTIs related to companion animal participants may be considered acceptable by members of the Victorian community (overall and by demographic subgroups).

Survey respondents reported their willingness to provide data and specimens as part of their hypothetical involvement in a HHTI. This will be described by the question, “Which of the following would you be comfortable to report or have collected in relation to your companion animal in a household transmission investigation (HHTI)?”, using a 5-point Likert scale ranging from “Very comfortable” to “Very uncomfortable”.

Survey respondents reported acceptability of self-collection and/or professional (vet) collection of swabs as part of their hypothetical involvement in a HHTI. This will be described by the question, “How acceptable are the following methods for the collection of nose and/or throat swabs from your companion animal in a HHTI?”, using a 5-point Likert scale ranging from “Very acceptable” to “Very unacceptable”

5. Describe ‘routine’ human-companion animal and companion animal-wildlife contact patterns (overall and by demographic subgroups).

Survey respondents were asked to report on their interactions with their ‘main’ companion animal and observed/potential interactions of their companion animal with wildlife. Interactions are described by the questions “How often do you personally...? How often does your companion animal...? Is your pet ever allowed to roam outside of the boundaries of your property by themselves? In the last 12 months, have you seen your pet interact with wildlife and/or other non-domesticated animals or their carcasses?” using Likert scale options “More than once a day, Once a day, More than once a week, Once a week, Less than once a week, Never” and other pre-defined responses to characterise interactions.

#### Secondary outcome measures

6. Assess survey completion rate and time taken, common stopping points (if applicable, overall and by demographic subgroups).

Internal data provided and/or derived from REDCap.

7. Describe participants’ perceptions about future pandemics ((if applicable, overall and by demographic subgroups).

Survey respondents were asked about their perceptions and concerns of future pandemics. Perceptions are described by the questions “How likely do you think another respiratory disease pandemic (like COVID-19) will occur in the next 10 years? What are your main concerns about having respiratory symptoms? If there was another respiratory disease pandemic (like COVID-19), how concerned do you think would you be about the following…?”, using a 5-point Likert scale ranging from “Very likely or very concerned to very unlikely or Not at all concerned”.

8. Describe participants’ perceptions about zoonoses (if applicable, overall and by demographic subgroups).

Survey respondents were asked about their knowledge of zoonotic disease risk. Knowledge is described by the questions “Do you think companion animals can get respiratory diseases from people? Do you think people can get respiratory diseases from companion animals?” using Yes/No answers, and free text fields.

9. Describe health care seeking behaviours and sources of trusted health information (if applicable, overall and by demographic subgroups).

Survey respondents were asked about how they access health information (source, frequency) or and their behaviours when sick. Outcomes are described by the questions “When you seek health advice and information, what do you look for? From which sources do you usually seek this information? Why do you choose the above sources of information? When you are sick or feeling unwell with respiratory symptoms, how often do you do the following?” using a 5-point Likert scale ranging from “Always to Never” and associated free text fields.

10. Describe health care seeking behaviours for companion animals (if applicable, overall and by demographic subgroups).

Survey respondents were asked about what they usually do first when their companion animal is sick by apparent severity and changes to interactions. Outcomes are described by the questions

“What signs would make you think that your companion animal might be unwell with an infectious disease? When your companion animal begins to show signs of mild illness, what do you usually do first? When your companion animal begins to show signs of severe illness, what do you usually do first?” using pre-defined options and associated free text fields.

11. Describe potential motivators for participation in HHTIs and supports required (if applicable, overall and by demographic subgroups).

Survey respondents were asked about several factors that may motivate or support them to take part in a HHTI. Outcomes are described by the questions “Which of the following factors would motivate you to take part in a HHTI? Including you, would anyone in your household require any of the following supports to participate in a HHTI?” using pre-defined options and associated free text fields.

### Section 3: Data cleaning and analysis

All data cleaning and analyses of quantitative data will be conducted in R statistical software version 4.3.1 (R Core Team 2023).

Responses will be included in the analysis dataset if consent was provided. Incomplete responses that only have mandatory demographic information such as age, gender, and no other information contributing to our outcomes will be excluded (may be fake responses due to earlier set up that allowed people to access prize draw survey without completing other components). Any other provided survey data will be analysed irrespective of survey completion status.

Missing data will be reported where appropriate and relevant to understanding of the question.

Quantitative data, including demographics and responses to multiple choice questions, will be summarised using descriptive statistics. Frequencies and proportions will be used to summarise categorical data. Household size will be summarised using the median and interquartile range.

Exploratory inferential statistical techniques (univariable multinomial and/or logistic regression models) will only be undertaken for all primary objectives to understand how survey responses (outcome/endpoints) vary among key demographic groups (see below). We will only report the estimate and 95% confidence interval for univariable analyses. We will not report p-values or conduct multivariable analyses.

The univariable analyses will only be performed if there are at least ten people in each outcome subcategory. Key subgroups per Table 1^#^ and below will be aggregated (where appropriate) such that these criteria are met.

- Gender: Male, Female, Non-binary
- Age: 18 – 29, 30 – 49, 50 – 64,  ≥65)
- Location: Metropolitan Melbourne vs regional/rural Victoria
- Languages other than English spoken at home: No, Yes
- Migration status: Born in Australia, Other country
- Medicare status: Eligible, Not eligible
- Education: Primary school, secondary school, vocational training, certificate/diploma/technical qualification, Bachelor’s degree, Post-graduate degree (e.g. Master’s, PhD)
- Current employment status: Employed, Not employed
- Children (<18) in Household: Yes, No
- Pets in Household: Yes, No

^#^Note these may be aggregated from data to reduce the number of categories explored. Univariable analyses will not be undertaken/reported where insufficient data exists (n=10) for a binary or categorical outcome.

Future work (depending on time and availability) may include a multiple correspondence analysis (MCA) to identify (visual) associations between categorical variables.

Qualitative data, such as free text responses to open-ended questions, may be thematically analysed if there is sufficient information (and they add further insights to multiple choice/matrix fields). This approach may help identify recurring themes, patterns, and perspectives related to participants’ KAP. Quotes will be retained for reporting (if deemed relevant).

Note: Similar ad-hoc analyses may be undertaken for the secondary objectives if there are differences for the primary objectives by subgroup. The team will consult prior to these being conducted.
